# Comparing Scientific Machine Learning with Population Pharmacokinetic and Classical Machine Learning Approaches for Prediction of Drug Concentrations

**DOI:** 10.1101/2024.05.06.24306555

**Authors:** Diego Valderrama, Olga Teplytska, Luca Marie Koltermann, Elena Trunz, Eduard Schmulenson, Achim Fritsch, Ulrich Jaehde, Holger Fröhlich

**Affiliations:** Department of Bioinformatics, Fraunhofer Institute for Algorithms and Scientific Computing (SCAI), Sankt Augustin, Germany; Department of Clinical Pharmacy, Institute of Pharmacy, University of Bonn, Bonn, Germany; Institute of Computer Science II, Visual Computing, University of Bonn, Bonn, Germany; Bonn-Aachen International Center for Information Technology (B-IT), University of Bonn, Bonn, Germany

**Keywords:** Scientific machine learning, Machine Learning, Oncology, Drug Therapy, Pharmacokinetics, Concentration Prediction

## Abstract

A variety of classical machine learning (ML) approaches have been developed over the past decade aiming to individualize drug dosages based on measured plasma concentrations. However, the interpretability of these models is challenging as they do not incorporate information on pharmacokinetic (PK) drug disposition. In this work we compare drug plasma concentraton predictions of well-known population PK (PopPK) modeling with classical machine learning models and a newly proposed scientific machine learning (MMPK-SciML) framework. MMPK-SciML lets us estimate PopPK parameters and their inter-individual variability (IIV) using multimodal covariate data of each patient and does not require assumptions about the underlying covariate relationships. A dataset of 541 fluorouracil (5FU) plasma concentrations as example for an intravenously administered drug and a dataset of 302 sunitinib and its active metabolite concentrations each, as example for an orally administered drug were used for analysis. Whereas classical machine learning models were not able to describe the data sufficiently, MMPK-SciML allowed us to obtain accurate drug plasma concentration predictions for test patients. In case of 5FU, goodness-of-fit shows that the MMPK-SciML approach predicts drug plasma concentrations more accurately than PopPK models. For sunitinib, we observed slightly less accurate drug concentration predictions compared to PopPK. Overall, MMPK-SciML has shown promising results and should therefore be further investigated as a valuable alternative to classical PopPK modeling, provided there is sufficient training data.

## 1. Introduction

During the last decade machine learning (ML) techniques have been increasingly employed for estimating drug plasma concentrations in dependency of pharmacokinetic (PK) parameters. Aside from concentration prediction, ML has also been used for other purposes in pharmacometric modeling, including data imputation, covariate selection and treatment response prediction. Thus, many authors have discussed in detail how ML can be used for different modeling approaches, such as population PK (PopPK), pharmacometric simulation, model-informed precision dosing and systems pharmacology to facilitate collaboration with computer scientists [1–4]. Main concerns regarding classical ML include i) lack of model interpretability and mechanistic insight, ii) difficulty to handle inter-individual variability (IIV), and iii) requirements of larger training data than typical in PopPK. However, some approaches have been proposed to address some of these limitations [5–9]. Lu et al. trained neural ordinary differential equations (ODEs) to predict PK profiles [8]. A combination of neural networks (NN) and knowledge-derived ODEs was employed by Qian et al. [10]. Similarly, Janssen et al. used a NN to learn covariate effects of drug concentrations [11]. We introduced PK-SciML [12], a Scientific Machine Learning (SciML) [13, 14] approach for learning an unknown absorption mechanism while simultaneously estimating PK parameters. However, our previous model was only evaluated on simulated data and only generates populational level predictions for different dose groups. Given that a SciML framework benefits from not requiring prior knowledge of the exact relationships between covariates and parameters while still incorporating domain expertise, we introduce a multimodal pharmacokinetic SciML (MMPK-SciML) approach. This extension of PK-SciML is designed to learn IIV based on multimodal covariate data, enabling the prediction of drug concentrations and simulation of complete concentration-time profiles. In this paper, we denote as multimodal a dataset incorporating different data modalities, including clinical measurements, demographic information, and other data types. As a case study, we use two real datasets for two different oncology treatments as examples for an intravenous (iv) and an oral treatment route and compare our model with different classical ML and PopPK models. We demonstrate that our model produces reliable predictions of drug plasma concentrations.

## 2. Methods

### 2.1. Data collection and preprocessing

#### 2.1.1. Fluorouracil (5FU)

In this work, plasma concentrations of patients who received fluorouracil (5FU)-based infusional chemotherapy at the Oncological Outpatient Clinic UnterEms in Leer, Germany, were retrospectively analyzed [15]. This study was approved by the local medical ethics committee, but trial registration was not conducted due to the retrospective nature. Plasma 5FU concentrations were obtained at steady-state during continuous infusion and quantified using the My5-FU™ immunoassay (Saladax Biomedical Inc., Bethlehem, PA, USA) [16]. The dataset included 549 concentration measurements from 157 patients and information on demographics, blood counts and adverse events. Doses were documented for all patients with their corresponding infusion times. Samples were drawn at steady-state 16.8-25.0 hours after start of infusion. All patients with documented therapeutic drug monitoring (TDM) of 5FU were included in the analysis, except one patient who only had one concentration below the lower limit of quantification (BLQ). Outliers were defined as samples with a concentration BLQ (<52 ng/mL) or a clearance above 1478 L/h (corresponding to 739 L/h/m² and a body surface area of 2 m^2^). This was deemed implausible due to reported ranges [17] and samples were excluded from the dataset. In total, we omitted eight entries from eight different patients (1.45% of all samples). For 5FU, one to nine samples per patient with a median of three were available for analysis. Weight was measured only once or twice per cycle and height only in the beginning of treatment. Thus, these values were assumed to remain unchanged until a new measurement was taken.

#### 2.1.2. Sunitinib

Sunitinib PK data were pooled from two PK/PD studies focusing on sunitinib treatment in patients with metastatic renal cell carcinoma (mRCC) and patients with metastatic colorectal cancer (mCRC) [18, 19]. The C-IV-001 study (EudraCT-No: 2012-001415-23, date of authorisation: 17.10.2012) was a phase IV PK/PD substudy of the non-interventional EuroTARGET project, which recruited patients with mRCC at nine medical centres in Germany and the Netherlands [18]. Sunitinib doses ranged from 37.5-50 mg daily, administered orally on a 4-week on/2-week off schedule. The C-II-005 study (EudraCT-No: 2008-00151537, date of authorisation: 11.06.2008) was conducted to investigate the beneficial effect of sunitinib added to biweekly folinate, fluorouracil and irinotecan in patients with mCRC and liver metastases. Patients were prescribed a daily dose of 37.5 mg sunitinib on a 4-week on/2-week off schedule taken orally [19]. Both studies were performed in accordance with the Declaration of Helsinki. A total of 308 sunitinib plasma and active metabolite (SU12662) concentrations were obtained from 26 mRCC and 21 mCRC patients [20]. Six sunitinib measurements BLQ (<0.06 ng/mL) from five different patients were excluded from the analysis, accounting to 1.95% of all samples. Times and dates of the respective doses were defined according to Diekstra et al. [20]. In the C-IV-001 study, up to 12 plasma samples were collected within three cycles during routine checkups. In the C-II-005 study, plasma samples were collected within two cycles at baseline, day two of each cycle, and afterwards approximately every second week, always before sunitinib intake [20]. For sunitinib, we had one to 14 samples per patient with a median of 6.5 in the dataset. In general, weight and height were only measured in the beginning of treatment; thus, these values were assumed to remain unchanged. Missing values were 12.9% for weight, 10.9% for height and 6.6% for body surface area (BSA). Notably, in some cases only BSA was reported, but not weight and height.

#### 2.1.3. Data preprocessing

The total datasets were split using a 10 times 5-fold cross-validation setting with a training-test split of 80/20, keeping data from one patient strictly in the same set to avoid a splitting bias. For the classical MLalgorithms, continuous features were scaled between zero and one.

### 2.2. Population pharmacokinetic modeling

For all PopPK analyses, we used the NONMEM^®^ version 7.5.0 and the PsN version 5.2.6. Pirana (version 3.0.0.) served as front interface. R version 4.3.1. was used in R Studio version 2023.06.1. The PopPK model for 5FU comprised of a one-compartment model with linear elimination to describe 5FU disposition [15]. While the 5FU clearance and its IIV were estimated, the volume of distribution and its IIV were fixed to previously estimated values [15] because they were mathematically (i.e. structurally) non-identifiable. The residual variability was modeled as proportional and the BSA centered on the population median, was included as a linear covariate on clearance. All available BSA values were used in modeling. Differently from the original model [15], the skeletal muscle index was not included as a covariate, because it was not available for all included patients. Schmulenson et al. used the first order conditional estimation with interaction (FOCE-I) method to estimate the parameters [15]. In addition, we employed stochastic approximation expectation maximisation with interaction (SAEM-I) to understand potential differences in parameter estimates and random effect distributions compared to FOCE-I [21].

Inter-occasion variability (IOV) was not included in the final model, because there was no significant improvement of the objective function value and the parameter precision by modeling IOV. First, we estimated the PK parameters for the patients in the training set using FOCE-I and SAEM-I and initial estimates based on reported values from Schmulenson et al. [15]. In the next step, the retrieved estimates were used to simulate the expected concentrations for the test data. Mean concentration values were calculated by subject from 1000 simulations without including residual variability (simulated IPRED) and without re-fitting the model. The structure of the PopPK model for sunitinib is shown in Eqs. 6-12. A two-compartment model for sunitinib disposition and a biphasic distribution for its active metabolite SU12662 were used [20, 22]. Presystemic formation of SU12662 was modeled via a hypothetical enzyme compartment incorporated into the central compartment of sunitinib. An intercompartmental clearance connected the central compartment and the enzyme compartment and was fixed to the liver blood flow. Furthermore, the fraction of sunitinib converted to SU12662 and the peripheral volume of distribution of sunitinib were fixed to reported values [20]. IIV was included for the central volumes of distribution for sunitinib and SU12662, the clearance of sunitinib and the fraction metabolized in a block matrix. Proportional errors for the parent drug and metabolite were used to describe the residual unexplained variability. For the PopPK model, missing weight data was imputed on the training data using the mean values for each sex according to Diekstra et al. [20]. After model fitting, we simulated the expected plasma concentrations for the patients in the test dataset.

### 2.3. Classical Machine Learning Algorithms

Various classical ML methods, including Random Forests, Gradient Boosting, Extreme Gradient Boosting (XGBoost), Light Gradient Boosting (LightGBM), Support Vector Machines (SVM) and simple NN with one and two hidden layers were used for concentration prediction in Python version 3.10. For 5FU the input variables consisted of dose, weight, lean body mass (LBM), fat mass (FM), BSA, age, sex, height and time since last dose. For sunitinib input variables comprised sex, age, weight, height, BSA and time since last dose. These potential covariates, despite most of them having been excluded in stepwise covariate modeling (SCM), were included to enable the ML algorithms to make use of potential previously missed relationships within the data as they have shown to outperform SCM in some cases [21]. Hyperparameter tuning was performed using the Bayesian hyperparameter optimisation framework Optuna (version 3.5.0) [22] and models were selected by applying 5-fold cross validation with the mean squared error as the objective function. Missing covariate data for sunitinib was imputed using a random forest approach (MissForest, version 2.4.2; missingpy, version 0.2.0) within the cross-validation process. The NNs were regularised applying common techniques such as drop-out, L1 regularization and gradient clipping to avoid overfitting.

To investigate whether the model performance of the classical ML methods could be further improved by adding synthetic data, the training dataset for sunitinib was augmented for each split according to Table S1. To simulate drug concentrations for each synthetic patient, we used the Diekstra et al. PopPK model fitted on training data within the cross-validation procedure.1000 synthetic patients with one measurement each were created within each cross-validation fold and added to the original training data. The consistency of the augmented with the real data can be seen in density plots for the covariates and goodness of fit plots for the concentrations in Figure S1.

### 2.4. Multimodal Pharmacokinetic SciML Model (MMPK-SciML)

The main motivation of MMPK-SciML was to overcome the limitations of PK-SciML [12], i.e., we wanted to build a model learning the IIV using neural networks and multimodal patient information. Following the classical PK framework, individual parameters using IIV are defined as follows:

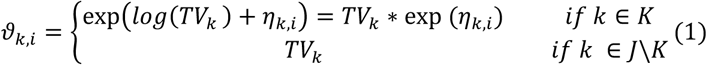

where *TV*_*k*_ is the typical population value of the parameter *k* ∈ *J*, and η_*k,i*_ (with *k* ∈ *K* ⊆ *J*) represents the IIV of that parameter for patient *i*. Notably, *J* is the total number of parameters, and *K* the subset of parameters with IIV. That means there can be parameters without IIV. Among those, a subset *L* ⊆ *J* is learned and the rest fixed.

Our proposed architecture is composed of 2 main blocks: i) a NN encoder which aims to predict the η values using patient covariates and ii) a structural well-defined ODE system to describe the PK dynamics. Therefore, given a total set of *J* patient parameters {*θ*_*k,i*_}_*k*∈ *J*_, a dose regimen, and a time horizon, the individual concentration profiles were predicted by solving the initial value problem of the ODE system.

Following PK-SciML [12] and Lu et al. [8] the dosage was added to the first compartment of the ODE system. Additionally, we fixed the initial conditions to zero to guarantee a plausible ODE system. Model implementation is available on GitHub at https://github.com/SCAI-BIO/MMPK-SciML.

#### 2.4.1. Variational Inference

Let *y*_*i,t*_, *t* ∈ τ denote the concentration profile measured at time points τ for patient *i*. Furthermore, *x*_*i*_ are patient-specific covariates. The mean μ and (log) variance log(σ^2^) of the approximate posterior distribution are learned from the observed data via an encoder neural network Φ_θ_:

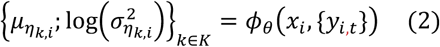

The initial value problem can then be solved by sampling from the distribution *N*(μ_η_, σ^2^_ni_) while taking advantage of the re-parametrization trick [23]. Specifically, the negative Evidence Lower Bound (ELBO) can be re-written as a loss function ℓ({*y*_*i,t*_}, {*y*_*i,t*_}):

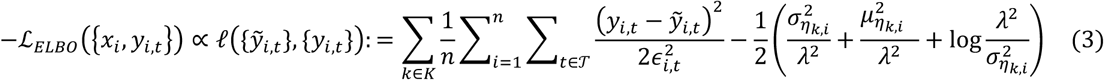

where є^2^_i,t_ is the variance of the measurement noise, λ a regularization parameter, and *y*°_*i,t*_ the ODE solution. For the following experiments we assumed a proportional error є^2^_i,t_ ∝ *y*_*i,t*_. More details can be found in Appendix A.

#### 2.4.2. Model details

##### 2.4.2.1. 5FU

Because all the measurements were taken at steady state, we considered them as conditionally independent and thus treated them as separate training samples. As structural ODE System we used an intravenous model as follows:

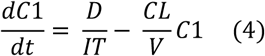

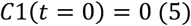

Where *D* is the dose, *IT* is the infusion time *CL* is the clearance, and *V* the volume of distribution.

To learn the random effects η, we defined Φ_θ_ as an encoder network using the concatenation of: the measured concentration, dose, weight, LBM, FM, BSA, age, sex and height was used as input for the first layer. Specifically, while {*TV*_*CL*_, *IIV*_*CL*_ } were estimated, *TV*_*V*_ = 46.1*L* was fixed. Figure 1 (top) shows an overview of our model architecture for 5FU. Model hyperparameters and more details can be found in Appendix A.

**Fig. 1.**
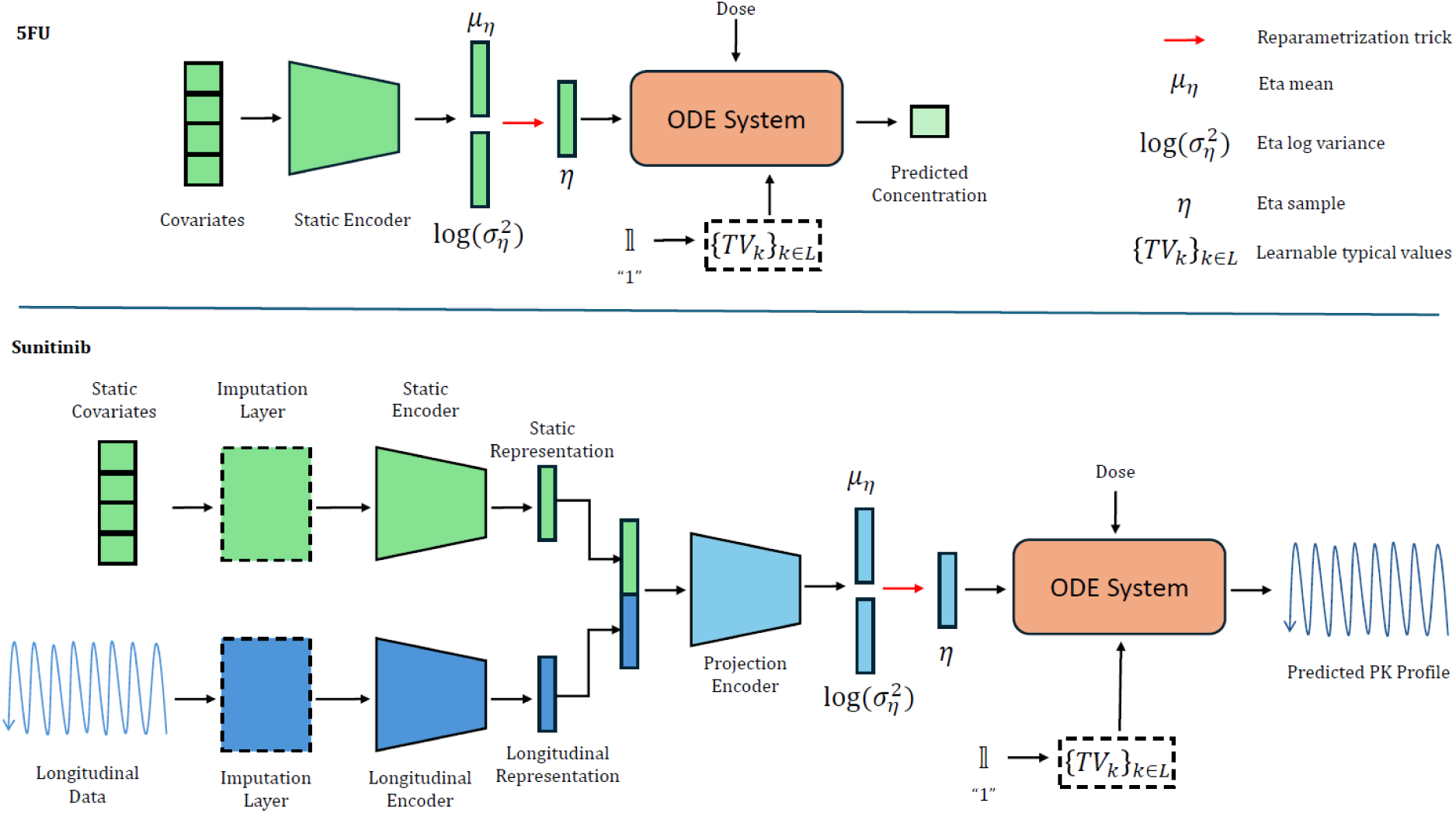
MMPK-SciML overview. The mean and log variance of the patient random effectś distribution is predicted with a neural network. At the same time, the population parameters are being learned and are used with an random effect sample to define the patient-specific parameters, which are used with the patient dose regimen to predict the PK profile.

##### 2.4.2.2. Sunitinib

We used as structural ODE system the model proposed by Diekstra et al. [20]:

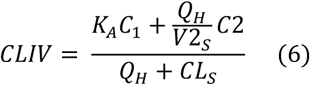

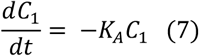

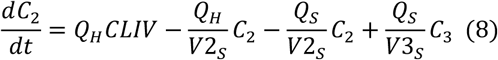

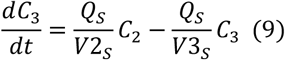

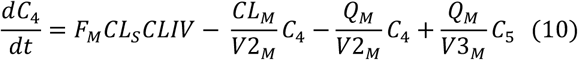

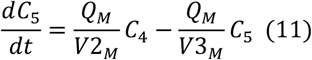

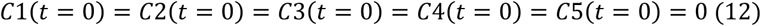

Where *K*_*A*_ is the absorption rate, *F*_*M*_ is the fraction metabolized to SU12662, (*CL*_*s*_, *Q*_*s*_), (*CL*_*M*_, *Q*_*M*_) are the clearance and intercompartmental clearance rate for sunitinib and its metabolite, respectively, *Q*_*H*_ is the liver blood flow, (*V*2_*s*_, *V*3_*s*_), (*V*2_*M*_, *V*3_*M*_) represent the volume of distribution and peripheral volume for sunitinib and its metabolite, respectively.

According to the original work by Diekstra et al. [20], the parameters of the sunitinib ODE system (eq. 5-11) should be scaled to make them comparable to literature values. Therefore, we first calculated the PK parameters following Eq (1), and then the values for *CL*_*s*_, *Q*_*s*_, *CL*_*M*_, *Q*_*M*_, *Q*_*H*_ were scaled by a factor of 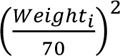 and those for *V*2, *V*3, *V*2, *V*3 by a factor of 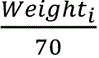.

Φ_θ_ was defined as a multimodal NN encoder containing three blocks. The first block was an encoder for static covariates {*x*_*i*_}. Since the sunitinib dataset includes measurements at multiple time points during the therapy cycle, the second block encoded the longitudinal covariates {*y*_*it*_ },for which we used the Time-LSTM [24] to capture the temporal dependencies. The output of both encoders was concatenated and used by a third block, the projection encoder, with 2 subnetworks each producing |*K*| outputs which define {μ_η_; log(σ^2^)}. We defined |*K*| = 4 corresponding to the IIV for *CL*, *V*2, *F*, *V*2. Population parameters {*TV*_*k*_ }_*k*∈*J*_ were either learned as part of the model training or fixed according to Diekstra et al. [20]. Figure 1 (bottom) shows an overview of our model architecture for sunitinib. Model hyperparameters and more details can be found in Appendix A.

### 2.5. Model Comparison

The goal of all algorithms was to predict single point plasma concentrations for patients in the test set based on information learned from the training data. To assess predictive performance, the mean absolute error (MAE), and the root mean squared error (RMSE) were calculated and compared for the different approaches used in this project. Goodness-of-fit (GOF) plots were used to support the quantitative results. In the case of the PopPK models, we used the fixed and random effect parameter estimates obtained on the training data to simulate individual predicted values for patients in the test dataset. Mean individual predictions were calculated from over 1000 simulations and compared against actual measurements. For the classical ML methods, the final predictions on the test dataset were used for the calculation of performance metrics. For the MMPK-SciML approach, the individual predictions for each patient were obtained using the means predicted by the encoder as random effects values because these represent the expected value.

To evaluate how well the models perform in simulating whole plasma PK profiles, prediction-corrected visual predictive checks (pcVPCs) were generated. These graphs could not be obtained for the classical machine learning approaches, because they are not generative.

## 3. Results

### 3.1. Dataset characteristics

A dataset of 541 fluorouracil (5FU) plasma concentrations from 156 patients as example for an IV administration and another dataset of 302 sunitinib and active metabolite concentrations each from 47 patients as example for a po administration were used for analysis. Baseline characteristics of all patients included in our analyses can be seen in Table 1.

**Table 1.** Baseline patient characteristics (median and range).

| 5 FU |  |  |
| --- | --- | --- |
| Demographics |  |  |
| Sex M/F | 96/60 |  |
| Age (years) | 64.5 (35-83) |  |
| Body surface area (m²) | 1.915 (1.35-2.85) |  |
| Therapy related details |  |  |
| 5FU dose (mg) | 4000 (2700-5720) |  |
| 5FU AUC (mg × h/L) <sup>a</sup> | 18.75 (8.1-92.3) |  |
| Therapy regimen |  |  |
| AIO <sup>b</sup> | 48 |  |
| FUFOX <sup>c</sup> (including monoclonal antibodies) | 41 |  |
| Paclitaxel/cisplatin/5FU/folate | 39 |  |
| Other | 28 |  |
| Tumor entity |  |  |
| Colorectal cancer | 79 |  |
| Gastroesophageal cancer | 48 |  |
| Pancreatic cancer | 16 |  |
| Other | 14 |  |
| Sunitinib |  |  |
|  | Patients with mRCC (*n* = 26) <sup>d</sup> | Patients with mCRC (*n* = 21) <sup>e</sup> |
| Demographics |  |  |
| Age | 64 (43–75) | 61 (33–85) |
| Sex M/F | 25/1 | 12/9 |
| Weight (kg) | 83 (65–106) | 73 (57–106) |
| Height (cm) | 180 (155–186) | 172 (149–184) |
| BMI (kg/m²) <sup>f</sup> | 25.7 (22.5–34.5) | 26.0 (13.3–39.3) |

**Notes:**
a Calculated by multiplying the infusion time with the measured steady-state concentration.
b Weekly 5FU infusion (2600 mg/m<sup>2</sup>) over 24 h in combination with folinate (500 mg/m<sup>2</sup>).
c Weekly 5FU infusion (2000 mg/m<sup>2</sup>) over 24 h in combination with folinate (500 mg/m<sup>2</sup>) and oxaliplatin (50 mg/m<sup>2</sup>).
d.e.f (abbreviations): mRCC, metastasized renal cell carcinoma; mCRC, metastasized colorectal cancer; BMI, body mass index.

### 3.2. Population pharmacokinetic modeling results

In the PopPK analyses, all PK parameters and their IIVs as defined in the original publications [15, 20] could be estimated for all data splits. The mean estimated parameter values were in a similar range to the originally estimated values for the whole datasets as depicted in Table 2 and the η values appeared to be normally distributed for all tested methods. There were no relevant differences between the estimated parameters and the simulated concentrations for the test data of the FOCE-I and SAEM-I methods.

**Table 2.**
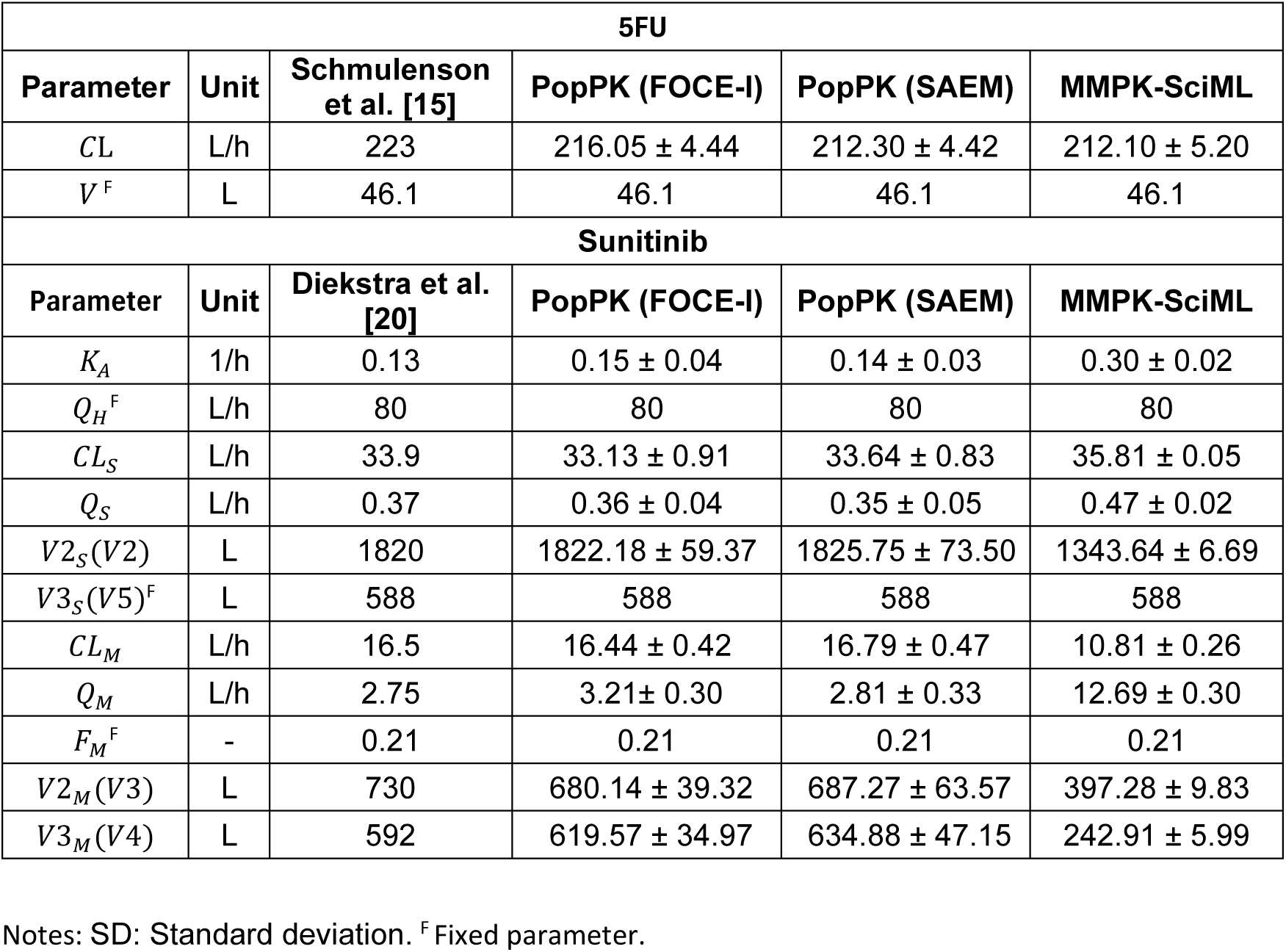
Cross validation population parameters (mean + SD) for 5FU and sunitinib.

For 5FU, using both FOCE-I and SAEM-I, we also observed normally distributed η. However, the GOF was still relatively poor, and showed wide confidence intervals(Figures 2, 4).

**Fig. 2.**
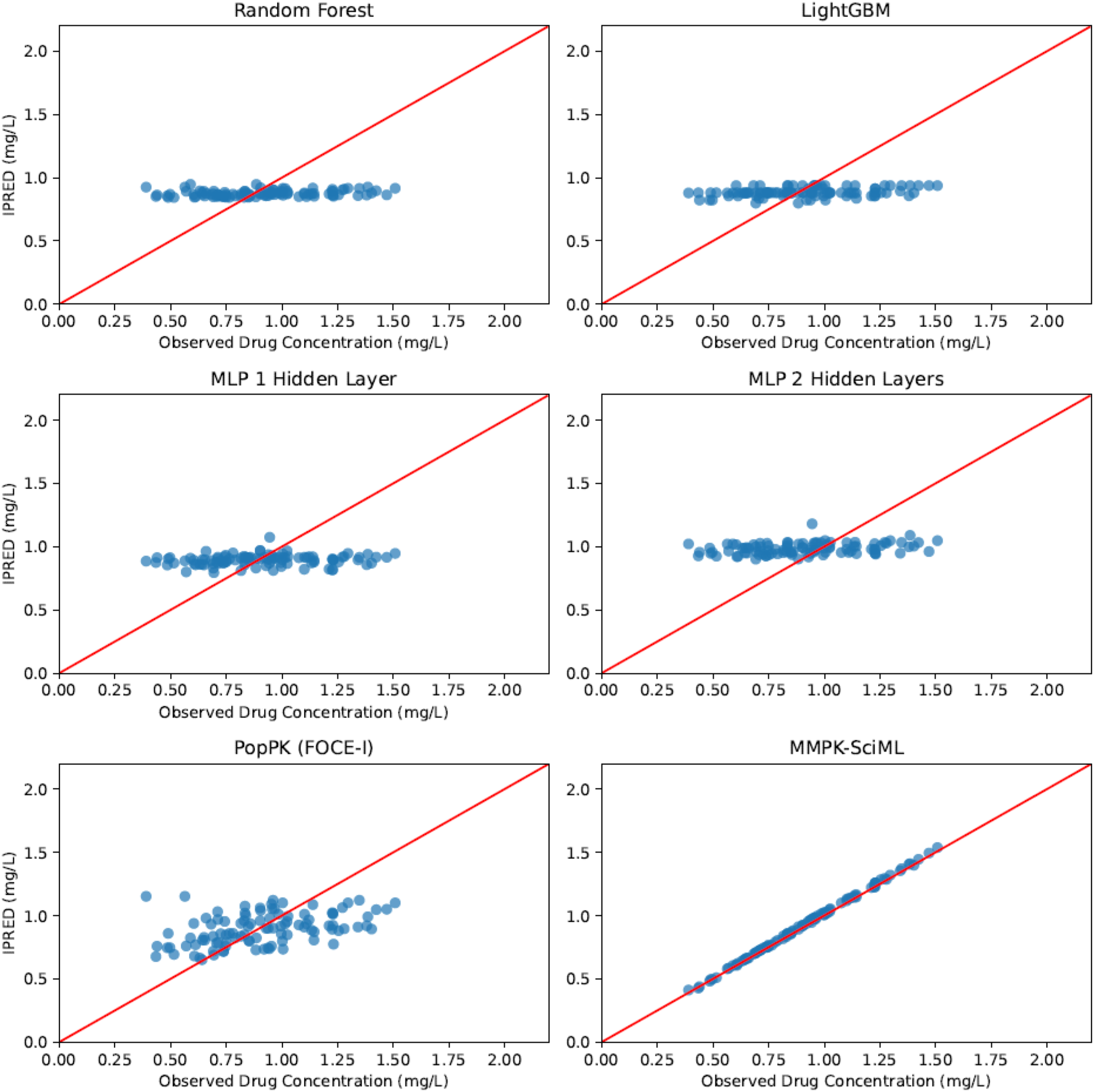
Goodness-of-fit (GOF) plots for the 5FU dataset showing predicted versus observed concentrations for selected trained models, with the results presented exclusively for the corresponding patients in the validation dataset.

In the case of sunitinib, convergence problems while fitting the PopPK model could only be solved by setting the initial estimates close to values reported by Diekstra et al. [20], yielding comparable results and good fits (Figures 3, 4). No significant differences between FOCE-I and SAEM-I fitting methods could be observed.

**Fig. 3.**
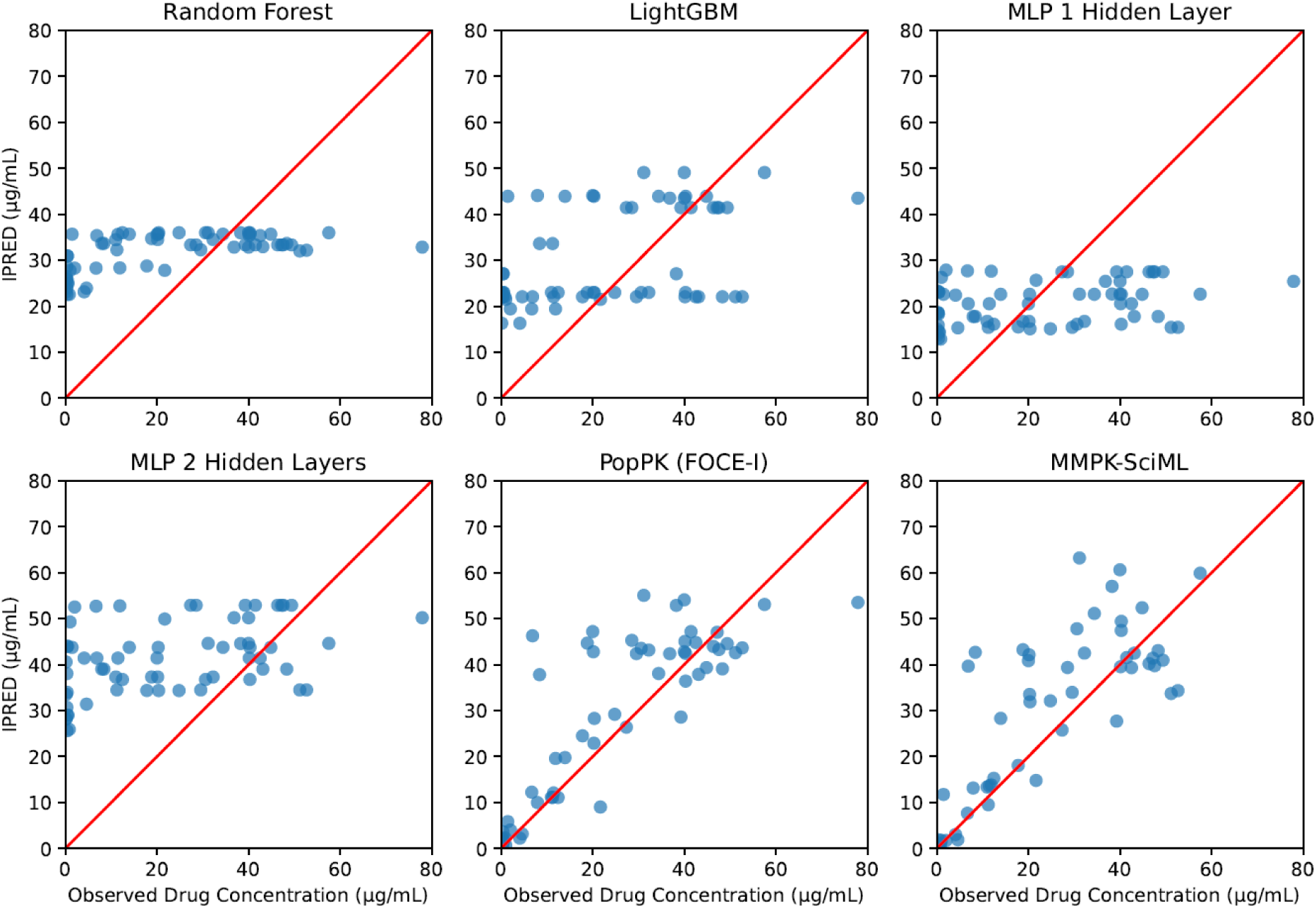
Goodness-of-fit (GOF) plots for the sunitinib dataset showing predicted versus observed concentrations for selected trained models, with the results presented exclusively for the corresponding patients in the validation dataset.

### 3.3. Classical machine learning methods

Optimized hyperparameters for all methods are reported in Supplementary Table S2. The proposed methods were not able to accurately predict plasma concentrations of both drugs as can be seen in the GOF plots in Figures 2 and 3, and the cross-validated accuracy metrics in Table 3. Augmentation of the original by synthetic data did not improve the situation. This can probably be attributed to the sparsity and high variability of the training data.

**Table 3.** Cross validation average metrics for 5FU and sunitinib.

| Model | 5FU |  | Sunitinib |  |
| --- | --- | --- | --- | --- |
|  | MAE | RMSE | MAE | RMSE |
| Random Forest | 0.23 ± 0.02 | 0.32 ± 0.12 | 18.39 ± 2.33<br>(18.50 ± 2.61) | 22.11 ± 3.13<br>(22.21 ± 3.15) |
| LightGBM | 0.23 ± 0.03 | 0.32 ± 0.12 | 16.81 ± 1.64<br>(19.23 ± 2.49) | 20.29 ± 1.93<br>(22.99 ± 3.02) |
| Multi-Layer Perceptron One Hidden Layer | 0.23 ± 0.02 | 0.32 ± 0.11 | 19.30 ± 2.63<br>(16.83 ± 2.52) | 23.71 ± 4.07<br>(21.27 ± 2.44) |
| Multi-Layer Perceptron Two Hidden Layers | 0.23 ± 0.02 | 0.32 ± 0.11 | 20.82 ± 3.31<br>(16.59 ± 3.10) | 24.96 ± 4.15<br>(21.49 ± 3.37) |
| PopPK (FOCE-I) | 0.22 ± 0.04 | 0.30 ± 0.12 | 9.69 ± 2.69 | 14.03 ± 3.91 |
| PopPK (SAEM-I) | 0.21 ± 0.03 | 0.28 ± 0.11 | **9.50 ± 2.52** | **13.69 ± 3.65** |
| MMPK-SciML | **0.04 ± 0.04** | **0.08 ± 0.12** | 12.55 ± 3.43 | 18.87 ± 5.12 |

Notes: We present the average value for the metrics (MAE: Mean Average Error, RMSE: Root Mean Square Error) and (±) the standard deviation across the 10-cross validation. In bold we show the best performance and in brackets the results with data augmentation. Metrics are reported when using sampling from the patient-specific distribution for test subjects.

### 3.4. MMPK-SciML

Our proposed MMPK-SciML model generated accurate predictions for both drugs. Figure 2, bottom row right, illustrates the GOF plots for 5FU. Opposed to classical ML methods a close correlation between the predictions and the real data was found. At the same time, cross-validated RMSE and MAE metrics were even lower than those of the PopPK model (Tables 3, S3). Especially for 5FU, we observed an at least five times lower MAE than all other methods, and at least 29% improvement of RMSE in all the cross-validation folds (Table S3).

Although the GOF plot of our MMPK-SciML model for sunitinib (Figure 3 bottom row right) was not as good as those for 5FU, our model still showed comparable performance to the PopPK methods, and better prediction accuracy than the best performing classical machine learning method LightGBM (Tables 3, S3). As can be seen in Figure 4, the MMPK-SciML models performed well in simulating test patients for both datasets, as the associated statistics of the real data are within the 90% confidence intervals (shaded region) of the predictions. Additionally, our models approximated the posterior η distributions in a reliable manner (Figure S2). Population parameters for all methods are reported in Table S4.

**Fig. 4.**
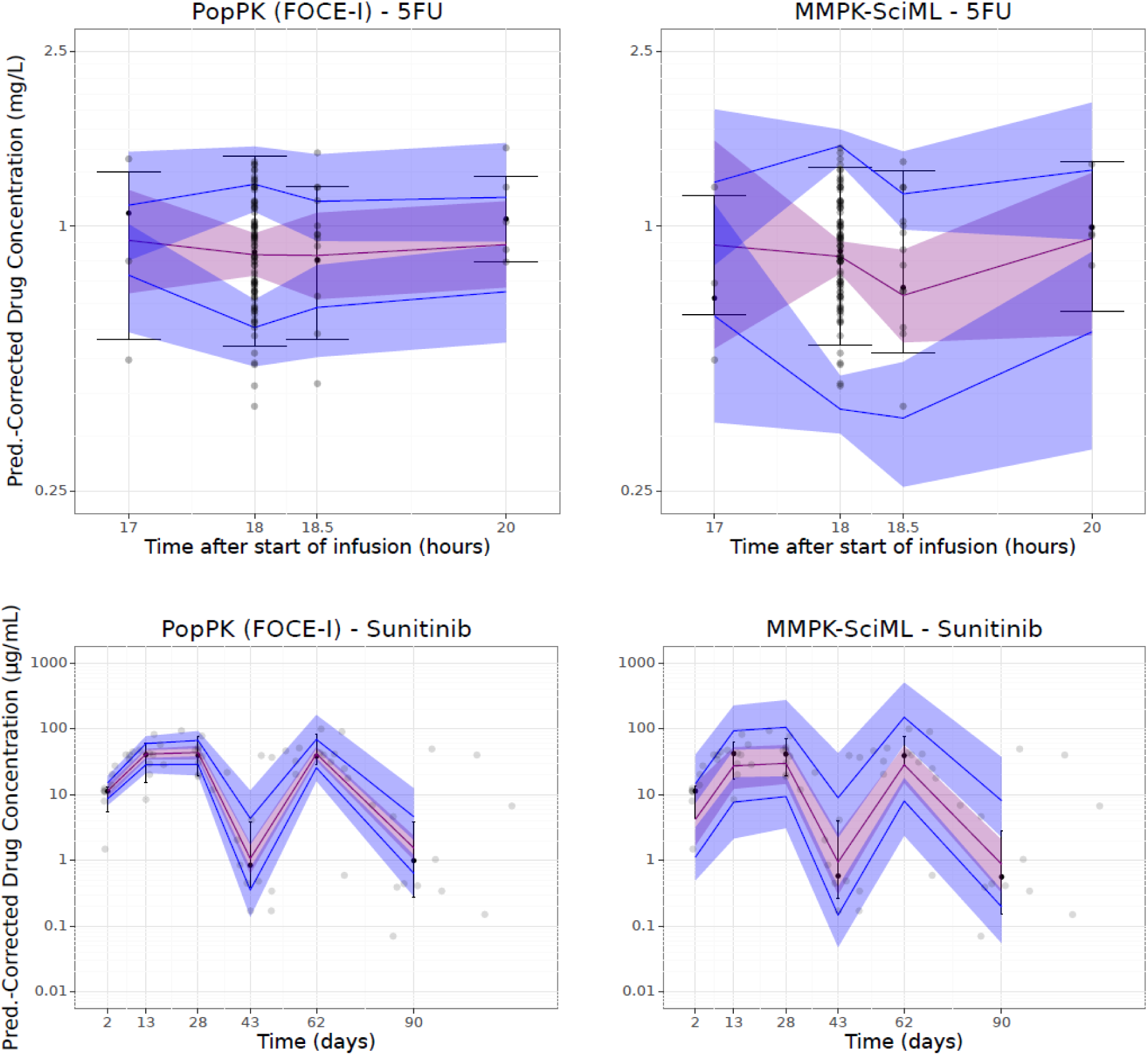
Prediction-corrected Visual Predictive Checks (pcVPC) plots for the 5FU (top) and the sunitinib (bottom) dataset.

MMPK-SciML has an implicit imputation mechanism. To better understand, how this may impact the comparison of model performances, we re-ran all models using data that was pre-imputed by MissForest. There were no significant differences from previously reported results.

## 4. Discussion

Our results demonstrate that generally a compartmental model structure is required to make accurate predictions of drug plasma concentrations, especially when measurements were not taken at steadystate such as in the case of sunitinib or when concentrations that are not trough are needed such as in the case of 5FU. Overall, only MMPK-SciML and PopPK methods were able to adequately describe the underlying drug disposition. In contrast to MMPK-SciML, which uses the same compartmental model structure as PopPK, classical ML models are entirely data-driven, lacking information about the concentration-time course and the time dependency between individual measurements. Without this structural guidance, classical ML algorithms cannot effectively learn key aspects of the data-generating process, whereas MMPK-SciML leverages problem-specific background knowledge to more accurately learn PK parameters. It should be noted that with classical ML extrapolation is generally infeasible. Since extrapolation is often required in practice, careful consideration is needed when applying classical ML models to data beyond their original training range. These limitations in addition to difficulties in model training when working with a small number of measurements and different dose schedules for more complex approaches [9, 27] were previously reported [12]. Data augmentation could not improve the performance of classical ML algorithms, suggesting that the inherent complexity of the temporal dynamics, the variance and the presence of concentrations that are far from the mean are difficult to learn by methods that have been designed for comparably simple tabular data only and use no information about the PK related processes. In this context it should be noted that covariate modeling techniques differ between the compared methods: While classical ML and MMPK-SciML implicitly model interactions of covariates, this is not the case for PopPK models. Here interactions have to be modeled explicitly, leading to a combinatorial explosion, especially if higher order (three-way, four-way) interactions are considered. Although any direct comparison between methods always remains limited due to the dependency on the data used, we altogether see a clear advantage of our proposed MMPK-SciML architecture in this regard. The model is particularly valuable in scenarios with many patient covariates where the influence of these covariates on random effects is not well understood, as it learns these relationships directly from the data. Furthermore, MMPK-SciML could be advantageous in cases where PK parameter (e.g. absorption) are challenging to estimate [12]. Since both treatment examples are rather complex (i.e., including dosing interruption or combination of different regimens), it could be insightful to apply the models to other treatment regimens to see how the performance differs in different setups. We leave this step as future work.

### 4.1. 5FU

In case of 5FU, GOF shows that the MMPK-SciML approach predicts drug plasma concentrations more accurately than PopPK models. MMPK-SciML predicted a similar population clearance to that obtained with PopPK models while achieving at least a three times lower MAE and an approximately 30% lower RSME (Tables S3, S4).The performance metrics of the classical ML approaches were in the range of the PopPK models, albeit with a worse GOF shown by the low correlation between the predictions and the real data (Figure 2, Table S3). However, a major limitation of our analyses was that the genotypes and the activity of the main metabolizing enzyme of 5FU, dihydropyrimidine dehydrogenase, which are important predictors for 5FU PK, were not available for our patient cohort. This information probably could have improved the performance of all tested models and should be reported in future studies.

### 4.2. Sunitinib

We observed relatively wide confidence intervals of the MMPK-SciML estimates. Although the absorption rate was predicted higher (0.13 1/h vs. 0.30 ± 0.02 1/h) and the central volume of distribution was predicted lower (1820 L vs. 1343.64 ± 6.69 L) compared to the original publication [20], the elimination and redistribution rates were similar across models in most of the cases (Table S4). Especially large differences (>40%) were observed in the estimates for the population parameters defining the concentration of the metabolite. Overall, the sunitinib analysis was more challenging than 5FU due to a rather small dataset composed of two different study populations increasing the variability. Moreover, more parameters had to be predicted due to the absorption process and the inclusion of metabolite concentrations, increasing the task complexity. However, considering that our model performed well despite these limitations (Figure 3, Table S3), we consider that our MMPK-SciML method would produce more narrow confidence intervals of parameter estimates if we had more training data, akin to 5FU.

## 5. Conclusions

This work highlights the need to use a structural model to effectively capture the time course of plasma concentrations in patients. In this regard we proposed a novel hybrid machine learning framework, which combines the flexibility of modern NN architectures with a compartmental model structure describing pharmacokinetic drug disposition. A limitation is the need for larger datasets compared to standard PopPK modeling approaches. On the other hand, our approach can capture inter-individual variability by learning patient-specific adjustments directly from the data, potentially bypassing the need for explicit covariate relationships. This offers an extension of traditional PopPK techniques and results in a simplification of the modeling process. Two possible directions of future research are i) to incorporate our model architecture into more complex frameworks for dosage adjustment, e.g. via reinforcement learning, ii) to develop methods for understanding the individual influence of covariates on model predictions.

## 6. Study Highlights

- **What is the current knowledge on the topic?** Machine Learning and Scientific Machine Learning (SciML) frameworks have shown promising results for pharmacokinetic modeling. However, methods for learning the inter-individual variability have not been widely investigated.
- **What question did this study address?** How well do population pharmacokinetic (PopPK) and classical machine learning (ML) approaches perform in comparison to a SciML approach for PK modeling? Can a neural network be employed in a SciML framework to learn inter-individual variability while making accurate PK predictions?
- **What does this study add to our knowledge?** The proposed MMPK-SciML model learns PopPK parameters and their inter-individual variability and may lead to more precise predictions than classical machine learning and PopPK approaches if enough data is given. Our proposed MMPK-SciML approach also addresses common drug development challenges such as missing values and different time sampling.
- **How might this change clinical pharmacology or translational science?** Our final framework provides an approach to learn patient-specific PK parameters and their IIV. Its potential for developing novel dosing strategies should be assessed in future studies.

## Data Availability

Data used in the present study could be available upon reasonable request to the authors

https://github.com/SCAI-BIO/MMPK-SciML

## Acknowledgements

We thank Ana Victoria Ponce Bobadilla for her input and explanations on how to create meaningful diagnostic plots for ML-enabled population PK models and Sven Stodtmann for the idea of encoding the concentration-time trajectory into parameters. We also thank Ana Socorro Rodríguez Báez for proofreading the manuscript and aiding with its structure. Furthermore, we thank Pharmetheus and especially consultant Anna McLaughlin for their student counselling and helpful explanations on the SAEM method.

## 7. Author contributions

U.J. and H.F. designed the research. D.V., O.T. and L.M.K. performed the research and analyzed the data., E.S. and A.F. developed the original population pharmacokinetic models, E.T. guided O.T. in coding the classical machine learning algorithms, and D.V., O.T., L.M.K., U.J. and H.F. wrote the manuscript.

### Use of AI

The tool DeepL Write was used by O.T. to refine the text. The tool ChatGPT was used by O.T. alongside Stack Overflow to assist in the coding process.

## Supplemental Information titles

TableS1.docx **Table S1.** Description of data augmentation for 5FU and sunitinib

TableS2.docx **Table S2.** Model hyperparameters for 5FU and sunitinib (Classic ML)

TableS3.docx **Table S3.** Full cross validation metrics for 5FU and sunitinib datasets

TableS4.docx **Table S4.** Full cross validation population parameters for 5FU and sunitinib datasets (PopPK and MMPK-SciML)

Fig S1.pdf **Fig. S1.** Density and goodness of fit plots to show compatibility of augmented data with real data (split 5)

FigS2.pdf **Fig. S2.** Posterior Random effects distributions for 5FU and sunitinib datasets (MMPK-SciML)

AppendixA.docx Technical details

## Notes

### Competing Interest Statement

H.F. received grants from UCB and AbbVie. The other authors declare no competing interest for this work.

### Funding Statement

This work was partially funded by Federal Ministry of Education and Research within the projects BNTrAinee (funding code 16DHBK1022).

### Author Declarations

The 5FU study (protocol-No: CESAR C-II-005, EudraCT-No; 2008-001515-37) with retrospective nature which was approved on 26.05.2008 by the ethics committee of the Albert-Ludwigs-Universitaet Freiburg, Freiburg, Germany. The C-IV-001 Sunitinib study (EudraCT-No: 2012-001415-23) was approved on 17.10.2012 by the ethics committee of the department of medicine of the Johann Wolfgang Goethe-Universitaet Frankfurt am Main, Frankfurt, Germany. It was a phase IV PK/PD substudy of the non-interventional EuroTARGET project, which recruited patients with mRCC at nine medical centres in Germany and the Netherlands

### Summary of Updates

Notation was clarified and possible uses cases where added

## References

1. Janssen A, Bennis FC, Mathôt RAA. Adoption of Machine Learning in Pharmacometrics: An Overview of Recent Implementations and Their Considerations. Pharmaceutics 2022; 14(9).

2. Stankevičiūtė K, Woillard J-B, Peck RW, Marquet P, van der Schaar M. Bridging the Worlds of Pharmacometrics and Machine Learning. Clin Pharmacokinet 2023; 62(11): 1551–65.

3. McComb M, Bies R, Ramanathan M. Machine learning in pharmacometrics: Opportunities and challenges. Br J Clin Pharmacol 2022; 88(4): 1482–99.

4. Talevi A, et al. Machine Learning in Drug Discovery and Development Part 1: A Primer. CPT Pharmacometrics Syst Pharmacol 2020; 9(3): 129–42.

5. Imai S, Takekuma Y, Miyai T, Sugawara M. A New Algorithm Optimized for Initial Dose Settings of Vancomycin Using Machine Learning. Biol Pharm Bull 2020; 43(1): 188–93.

6. Hu PJ-H, Wei C-P, Cheng T-H, Chen J-X. Predicting adequacy of vancomycin regimens: A learning-based classification approach to improving clinical decision making. Decision Support Systems 2007; 43(4): 1226–41.

7. Tang J, et al. Application of Machine-Learning Models to Predict Tacrolimus Stable Dose in Renal Transplant Recipients. Sci Rep 2017; 7: 42192.

8. Lu J, Deng K, Zhang X, Liu G, Guan Y. Neural-ODE for pharmacokinetics modeling and its advantage to alternative machine learning models in predicting new dosing regimens. iScience 2021; 24(7): 102804.

9. You Dubout W. An Algorithmic Approach to Personalized Drug Concentration Predictions. Lausanne, EPFL; 2014.

10. Qian Z, Zame W, Fleuren L, Elbers P, van der Schaar M. Integrating Expert ODEs into Neural ODEs: Pharmacology and Disease Progression. In: Advances in Neural Information Processing Systems. Integrating Expert ODEs into Neural ODEs: Pharmacology and Disease Progression; 2021. Curran Associates, Inc; 11364–83.

11. Janssen A, Leebeek FWG, Cnossen MH, Mathôt RAA, the OPTI-CLOT study group, SYMPHONY consortium. Deep compartment models: A deep learning approach for the reliable prediction of time-series data in pharmacokinetic modeling. CPT Pharmacometrics Syst Pharmacol 2022; 11(7): 934–45.

12. Valderrama D, Ponce-Bobadilla AV, Mensing S, Fröhlich H, Stodtmann S. Integrating machine learning with pharmacokinetic models: Benefits of scientific machine learning in adding neural networks components to existing PK models. CPT Pharmacometrics Syst Pharmacol 2024; 13(1): 41–53.

13. Baker N, et al. Workshop Report on Basic Research Needs for Scientific Machine Learning: Core Technologies for Artificial Intelligence; 2019.

14. Rackauckas C, Ma Y, Martensen J, et al. Universal Differential Equations for Scientific Machine Learning; 2020, arXiv preprint. http://arxiv.org/pdf/2001.04385.pdf2001.04385v4.

15. Schmulenson E, Zimmermann N, Müller L, Kapsa S, Sihinevich I, Jaehde U. Influence of the skeletal muscle index on pharmacokinetics and toxicity of fluorouracil. Cancer Med 2023; 12(3): 2580–9.

16. Beumer JH, et al. Multicenter evaluation of a novel nanoparticle immunoassay for 5-fluorouracil on the Olympus AU400 analyzer. Ther Drug Monit 2009; 31(6): 688–94.

17. Milano G, et al. Influence of sex and age on fluorouracil clearance. J Clin Oncol 1992; 10(7): 1171–5.

18. van der Zanden LFM, et al. Description of the EuroTARGET cohort: A European collaborative project on Tar-geted therapy in renal cell cancer-GEnetic- and tumor-related biomarkers for response and toxicity. Urol Oncol 2017; 35(8): 529.e9–529.e16.

19. Mross K, et al. FOLFIRI and sunitinib as first-line treatment in metastatic colorectal cancer patients with liver metastases--a CESAR phase II study including pharmacokinetic, biomarker, and imaging data. Int J Clin Pharmacol Ther 2014; 52(8): 642–52.

20. Diekstra MH, et al. Population Modeling Integrating Pharmacokinetics, Pharmacodynamics, Pharmacogenetics, and Clinical Outcome in Patients With Sunitinib-Treated Cancer. CPT Pharmacometrics Syst Pharmacol 2017; 6(9): 604–13.

21. Sibieude E, Khandelwal A, Girard P, Hesthaven JS, Terranova N. Population pharmacokinetic model selection assisted by machine learning. J Pharmacokinet Pharmacodyn 2022; 49(2): 257–70.

22. Akiba T, Sano S, Yanase T, Ohta T, Koyama M. Optuna. In: Proceedings of the 25th ACM SIGKDD International Conference on Knowledge Discovery & Data Mining. Optuna; 2019. New York, NY, United States: Association for Computing Machinery; 2623–31.

23. Kingma DP, Welling M. Auto-Encoding Variational Bayes; 2013, arXiv preprint. http://arxiv.org/pdf/1312.61141312.6114v11.

24. Baytas IM, Xiao C, Zhang X, Wang F, Jain AK, Zhou J. Patient Subtyping via Time-Aware LSTM Networks. In: Proceedings of the 23rd ACM SIGKDD International Conference on Knowledge Discovery and Data Mining. Patient Subtyping via Time-Aware LSTM Networks; 2017. New York, NY: ACM; 65–74.

